# RubricOE: a learning framework for genetic epidemiology

**DOI:** 10.1101/2021.03.09.21253105

**Authors:** Subrata Saha, Aldo Guzmán-Sáenz, Aritra Bose, Filippo Utro, Daniel E. Platt, Laxmi Parida

## Abstract

Genetic epidemiology is a growing area of interest in the past years due to the availability of genetic data with the decreasing cost of sequencing. Machine learning (ML) algorithms can be a very useful tool to study the genetic factors on disease incidence or on different traits characterizing a population. There are many challenges that plagues the field of genetic epidemiology including the unbalanced case-control data sets, fallibility of standard genome wide association studies with single marker analysis, heavily underdetermined systems with millions of markers in contrast of a few thousands of samples, to name a few. Ensemble ML methods can be a very useful tool to tackle many of these challenges and thus we propose RubricOE, a pipeline of ML algorithms with error bar computations to obtain interpretable genetic and non-genetic features from genomic or transcriptomic data combined with clinical factors in the form of electronic health records. RubricOE is shown to be robust in simulation studies, detecting true associations with traits of interest in arbitrarily structured multi-ethnic populations.

## Introduction

In recent years, with the availability of large cohorts for complex diseases, identifying the associated genetic variants as well as gene-environment interactions has become more challenging. Genome-wide association studies (GWAS) detect associations between genetic variants (single nucleotide polymorphism or SNP) and traits is a powerful tool but is sensitive to sample sizes, population structure, rare variants, environmental factors, etc. However, GWAS has several challenges in disentangling environmental effect from true genetic associations of a disorder leading to spurious associations. It also suffers from the curse of dimensionality with the availability of millions of SNPs for several thousands individuals, leading to the “small *n*, large *p* problems” (due to the under-determined nature of the genotype matrix). With the availability of more population controls than cases for binary traits due to low prevalence of many conditions/diseases the genotype data sets are often unbalanced. The scale of the data as well as the unbalanced nature of binary traits pose a substantial challenge for GWAS in large cohorts [1]. The SNPs are often genotyped with linkage disequilibrium (LD) between them resulting in correlated variables, which needs to be taken into account to find true genetic associations. Therefore, standard multivariable statistical approaches like linear or logistic regression are not well suited for genome-wide data [2]. Due to these shortcomings there has been a need for sophisticated machine learning approaches.

Machine learning (ML) algorithms can learn from the data without making any model assumptions. It provides several alternatives for performing multi-SNP analyses away from the single marker analysis as done in GWAS. It can employ techniques such as regularization and cross-validation in regression to tackle overfitting issues for under-determined data sets. Machine learning methods have been applied on a broad range of problems in genomics including how to recognize locations of transcriptions start sites, promoters, enhancers, etc in a genome sequence [3]. It has also been employed for genomic selection in plant breeding [4] as well as in GWAS [2] and genetic epidemiology [5]. ML can also elucidate non-linear SNP-SNP epistatic interactions, such as testing for *N* ^2^ SNP-SNP combinations for *N* SNPs. Generally, with multiple hypothesis Bonferroni corrected thresholds, this can be a difficult task with linear regression based techniques. For binary traits of complex diseases, the challenge is to identify clusters of alleles that suggest interactive pathways which are potential therapeutic targets.

In order to do that, it is important to understand how error analysis and statistical power interact in ML as probes to characterize identified candidates for pathogenic SNPs or other features. Therefore, we present a ML pipeline, called RubricOE, namely, a rubric for multi-omics association studies and genetic epidemiology. It includes main stages for feature selection, and for cross-validation applied to the selected feature set from the first stage. We consider the effect of error analysis to feature selection and cross validation. Lastly, it also considers the possibility of identifying SNPs that are purely epistatic. We find that RubricOE can correctly identify true associations in simulated data. It observes a trade-off with genetic effect in simulated datasets leading to minimal spurious associations for increased genetic effect and vice versa.

RubricOE is a robust ML pipeline that ranks disease associated SNPs and predicts the outcome of different disorder or traits. It provides further interpretation of the selected associations with their ranks and scores for studying their relative importance. The ranks and scores of SNPs closely imitates Polygenic Risk Scores (PRS) and thus can be a very useful tool for studying heritability of different traits. RubricOE is less sensitive to noise than standard GWAS and provides different optimizations in selecting hyper-parameters further elucidating the importance of each selected SNP. It also provides a framework to integrate clinical Electronic Health Record (EHR) information along with genomic data, providing further information on interactions between genetic and non-genetic factors associated with a disease or trait.

## Results

### Simulated Data

We applied RubricOE on six different simulation data sets with 1,000 individuals across 10,000 SNPs, related to two different phenotype distribution cases for each of the three population structure simulation scenarios related to genotypes. We observe that in the most challenging scenario with very low genetic effect (10%) consisting of only ten causal SNPs, RubricOE detects 30% of causal SNPs in the real-world TGP scenario. The Youden’s J statistic on the validation data in this case is understandably very low due to the overwhelming amount of noise in the dataset. Alternatively, when the genetic effect (70%) is substantially more than environmental effect and noise, RubricOE is able to discover accurately around 90% of the true causal SNPs in PSD and 70% in TGP, respectively. The Youden’s J statistic are also relatively higher with around 0.45 in PSD model, even though there are only ten true causal SNPs contributing to the binary phenotype. The score curves of the above experiments show that the peak scores of each feature in RubricOE is reached by using a very small set of features. As RubricOE computes the rank of the features by the descending order of their score and then computes Youden’s J statistic iteratively on the dataset, it gives an opportunity to understand how much a subset of feature contributes to the total classification accuracy of the learning algorithm. Thus, we observe, in case of PSD which accurately identifies 90% of causal associations, it needs only the first few hundred features to reach its peak cumulative score (Figures 4-6 in Appendix), deeming the rest of the SNPs non-informative. We see similar patterns in other simulation scenarios such as TGP and BN as well. We can leverage this information in making RubricOE faster by asking it to compute curves only with a few thousand features instead of the large number of SNPs, genes and clinical features, sometimes amounting to a few millions. This provides a significant scale-up in computational time of the pipeline. As SVM has quadratic complexity, this feature of RubricOE is particularly useful.

**Table 1:**
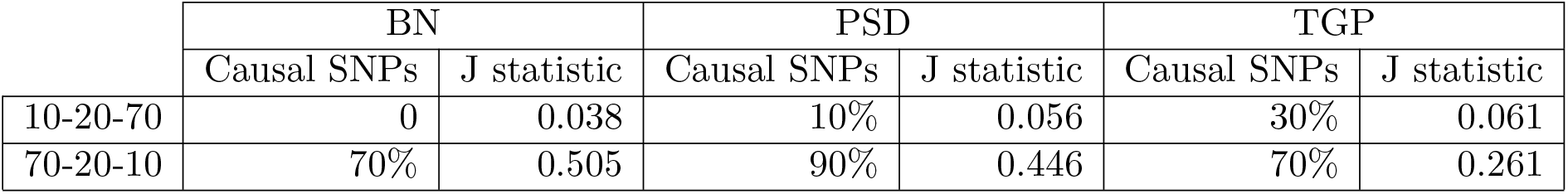
RubricOE performance on validation data in simulation studies evaluated by the percentage of causal SNPs detected and the mean Youden’s J statistic observed.

## Discussion

RubricOE provides a ML framework with which a stable set of features discriminating between the healthy controls and diseased patients, can be obtained. This differs from a single marker analysis as performed in GWAS. ML algorithms allow multi-SNP analysis along with interpretation of the subset of SNPs classifying different diseases. It ranks the SNPs along with their relative scores as calculated by Youden’s J statistic. One of the advantages of ML algorithms is in applying criteria for accepting features that focus on aggregated predictive power. For regression, the fraction of variation predicted by a feature is closely related, and for standard regression equal, to the square of the z-score for the coefficient. The downside is that large models may be underdetermined, and feature selection may identify false features due to overfitting. Regulation (Lasso and Ridge) mitigates that, but complicates the relationship between feature significance and how much variation is accounted for by the feature. PRS is comprised by most of the features of ML algorithms explored here, including susceptibility to overfitting. So the diffusion of functional associations may be spurious.

Associations can be spurious due to environmental effects as well as genotyping errors, etc. We explored robustness of RubricOE to such effects in the simulation studies and observed that the framework was able to correctly detect about 70% of true causal associations in the simulation scenarios closely emulating real-world population structure when the genetic effect is significantly more than non-genetic factors. RubricOE is also able to detect about 20-30% of causal associations when the genetic effect is only 10%, among which only 0.1% is causal. This demonstrates the prowess of RubricOE in detecting rare associations in the presence of noise.

The estimate of an individual’s genetic liability to a trait or disease of interest is captured by the PRS computation. It returns a score which is parallel to the Youden’s J statistic described here. Another factor that RubricOE can provide a solution to is the infamous “missing heritability” issue. Single marker analysis as done in GWAS does not account for the heritabiity of a trait or disease across generations. PRS aims to solve that problem, but, RubricOE provides a straight-forward one-pass solution for doing multi-SNP tests. One of the advantages of ML algorithms is in applying criteria for accepting features that focus on aggregated predictive power. For regression, the fraction of variation predicted by a feature is closely related, and for standard regression equal, to the square of the z-score for the coefficient. The downside is that large models may be underdetermined, and feature selection may identify false features due to overfitting. Regularization (Lasso and Ridge) mitigates that, but complicates the relationship between feature significance and how much variation is accounted for by the feature. We showed that, for random sampling variation, cross-validation does not really boost power. However, cross-validation is an effective way to recognize biased variations (e.g. batch effects). PRS is comprised by most of the features of machine learning algorithms explored here, including susceptibility to overfitting. So the diffusion of functional associations may be spurious. A detailed study on the parallels between PRS and Youden’s J statistic as well as demonstrate how it approaches the “missing heritability” issue across multi-ethnic cohorts would highlight the importance of studying other ML algorithms such as neural network or deep learning in disorder trait prediction and interpretable feature selection. RubricOE, or a rubric for omics and epidemiology can be useful in stable feature selection from integrated multi-omic and clinical data, elucidating their interactions.

## Methods

### Datasets

#### Simulation Study

We generated an extensive set of simulations with real-world challenging scenarios to demonstrate the robustness to different scenario and power to detect true disorder or trait associated SNPs. We simulated and analyzed six different data sets with 1,000 individuals and 10,0000 SNPs, pertaining to three population structure scenarios and for each, two phenotype simulation scenarios. The data was simulated based on a binary trait model using the Odds Ratio (OR) as the classifier for disease status from a quantitative trait model described in previous work [6].

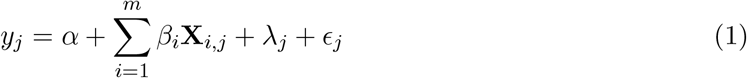

where *β*_*i*_ is the genetic effect of SNP *i* on the trait, *λ*_*j*_ is the random non-genetic effect and *ϵ*_*j*_ is the random noise variation for individual *j*. **X**_*i,j*_ is the *i*^*th*^ marker for the *j*^*th*^ individual and *y* ∈ ℝ^*m*^ is the trait response variable (binary or continuous).

For the genotype data, we simulated allele frequencies using (i) Balding-Nichols (BN) model [7] based on allele-frequency and *F*_*ST*_ estimates calculated on the HapMap data set, (ii) three different levels of admixture by varying the parameter *α* from {0.01,0.1,0.5} in Pritchard-Stephens-Donnelly model (PSD) [8] and (iii) structure estimated from 1000 Genomes Project (TGP) [9]. The three scenarios allow us to simulate genotypes with varying degree of population struc-ture [6, 10]. BN provides three arbitrarily structured group of populations who are not mixing with each other. This provides an ideal case of structure to evaluate the model. PSD model accounts for admixture between these arbitrarily structured clusters of individuals (such as individuals between each group share alleles between them) and the degree of admixture can be altered by the parameter *α*. Lastly, TGP model provides the most real-world like scenario where the individuals are sampled from allele frequencies corresponding to different population across the world as represented in the 1000 Genomes data set [9].

We used different variance ratio for the phenotype simulation scenarios. For the first scenario, we allowed 70% genetic effect, 20% environmental effect and 10% noise effect. This allowed us to study whether the model can accurately detect the true associations when the genetic effect is substantial. In comparison, we reversed the variances of the genetic effect (10%) and noise (70%) to simulate a more challenging scenario. The causal genetic effect is simulated by independently simulating the first 0.001 of the total number of SNPs, *n* (10,000 SNPs). *β*_*i*_ *Normal*(0, 0.5) for *i* = 1, 2, …, 0.001 * *n*. For all other *i* > 0.001 * *n* we set *β*_*i*_ = 0.

#### RubricOE

The ML pipeline,”RubricOE” outlined in Figure 2, has 3 major components: Quality Control (QC) of input omics data, Iterative Feature Selection (IFS), and Stable Set Construction (SSC). Optional selection of machine learning components with hyper-parameters may be employed in each to optimize performance. The pipeline employs a nested test-training set configuration. The outer test-training set split reserves the test set (“validation”) for final SNP evaluation and denotes the training set as “working” data. Within the latter, further train-test splits are performed to rank the features.

**Figure 1:**
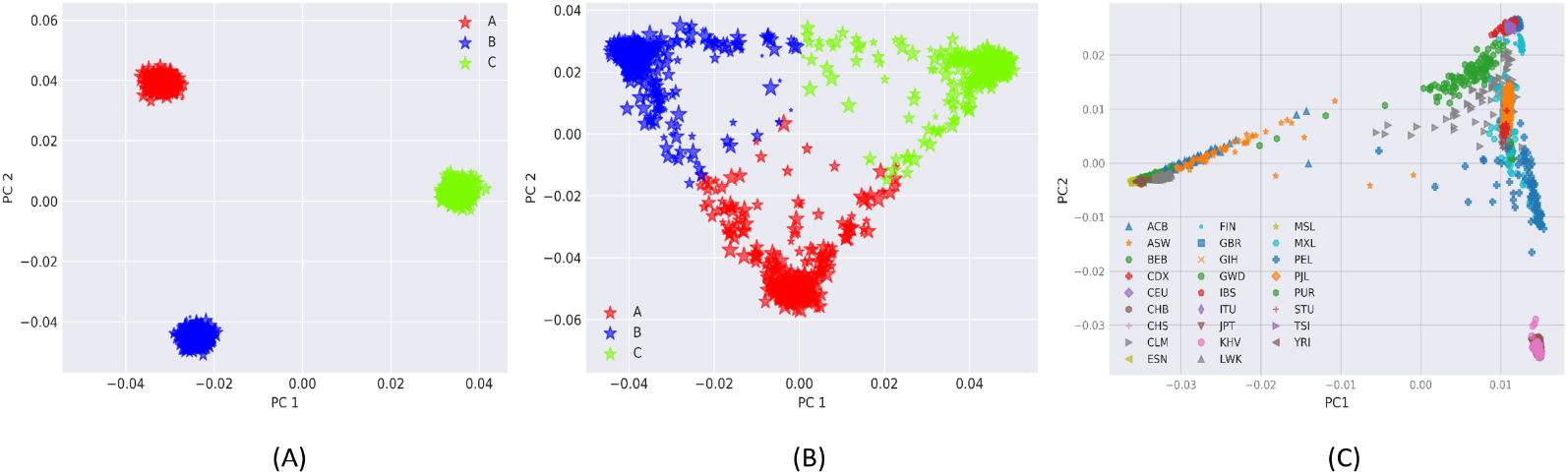
Projection of the samples from three populations simulated from (i) BN (ii) PSD (*α* = {0.1, 0.1, 0.1}) and (iii) TGP model on the top two axes of variation.

**Figure 2:**
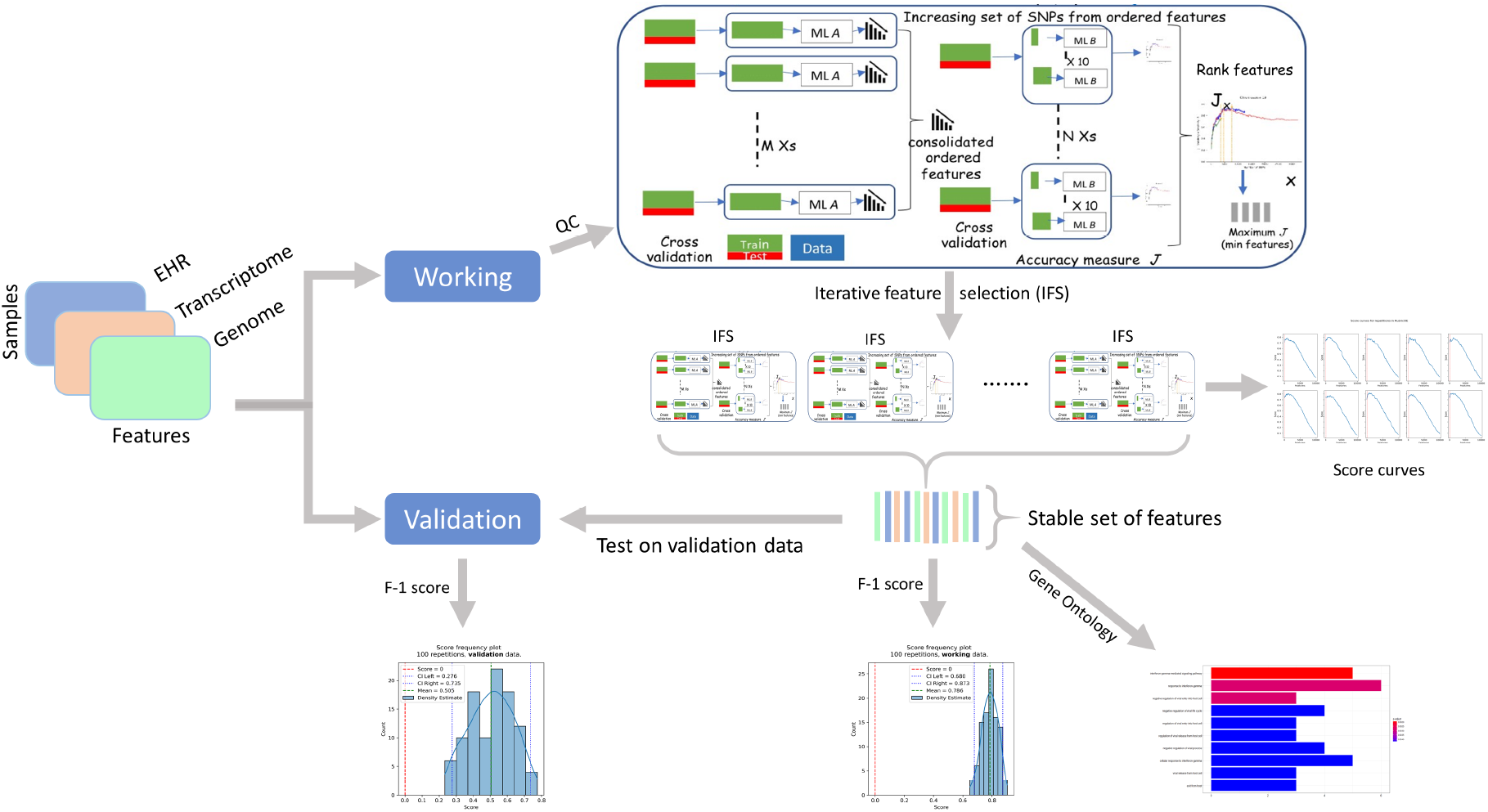
Components of RubricOE, the ensemble ML pipeline producing a stable set of features from multi-omics data.

The working data is used within the IFS stage. In this stage, replications of data test-training splits are each subject to iterative feature ranking, using machine learning algorithms (Ridge Regression in this case), and scored against the replicated test sets using a scoring metric (e.g. Youden or *h*^2^) using another machine learning algorithm (Support Vector Machines or SVM with linear kernel in this case) to identify an subset of SNPs. If the ML is non-linear in character, feature selection is based on recursive feature elimination (RFE) [11] is an option we have used. These rankings are combined to form a final candidate set. Models constructed from the final set are applied to the “unseen” (test) set for final scoring. Iterative replications are performed subsequently, and the set of SNPs that persistently survive in the intersection are called the “Stable Set”.

#### Iterative Feature Selection

##### Robust SNP ranking

Some of the ranking algorithms that were applied here, besides application of simple ridge regression, are described here. We attempt to make our SNP ranking model robust by employing multiple linear SVMs (LSVMs) in a concerted way. Each LSVM runs on slightly different data sets randomly sampled from original dataset *D*. In the experiment, we randomly picked 90% unique samples from *D* for each LSVM to build a robust learning model. Due to this small change, the weight associated with each SNP is not identical for different runs. By averaging the weights across a set of weight vectors from LSVMs of the same configuration, we can make the weight vector robust. Since weights are directly associated with the importance of the SNPs, ranking should also be robust.

Let the number of LSVMs to rank a set *S* of SNPs of interest be *L*. Each SVM *l* ∈ *L* is trained on a slightly smaller dataset *D*′ to build an inductive model and we get a corresponding weight vector *w*_*l*_ (1 ≤ *l* ≤ |*L*|). According to [11], the significance of the *i*^*th*^ SNP 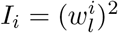 where 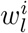 represents the *i*^*th*^ weight component of 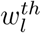 weight vector. At the end of this step, we get a set of |*L*| weight vectors. Since, in each run, we introduce slightly different dataset *D*′ by randomly sampling the original dataset *D*, the weight vectors will be different from each other. To make it stable we normalize each weight vector and average each component. Each weight vector *w*_*l*_ will have *n* components and is normalized as follows: 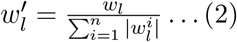. The *i*^*th*^ component of the final weight vector *W* is formed as follows: 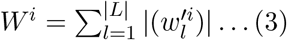 Now, the importance of the *i*^*th*^ SNP is defined as 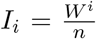. We sort the SNPs based on their significance in non-decreasing order. So, the highest significant SNP will have lowest rank, the 2^*nd*^ highest significant SNP will have 2^*nd*^ lowest rank, and so forth.

##### Robust SNP subset selection

Finding optimal set of features from *n* features will take 𝒪(2^*n*^) time which is exponential in the number of features. To reduce the search space, we follow two step procedure. At first, we employ robust SNP ranking algorithm (as described above) to rank SNPs based on their significance. In the second step, we linearly search through the ranked SNP space to get the subset of SNPs maximizing the classification accuracy (such as Youden index). Next, we describe the procedure in detail.

Let *r*_0_, *r*_1_, *r*_2_, …, *r*_*n−*1_ be the ranks of the given SNPs where *r*_0_ is the smallest rank, *r*_1_ is the second smallest rank, etc. Next, we take top *x* SNPs (i.e. *r*_0_, *r*_1_, …, *r*_*x*_ where *x* « *n*) and compute Youden index using 10-*fold* cross validation based on those *x* SNPs. Subsequently, we compute Youden indices based on top 2*x* SNPs, top 3*x* SNPs, etc. As soon as there is no improvement over the maximum Youden index seen so far in succeeding iterations, we stop the linear search and return back those SNPs having maximum Youden index. These SNPs constitute our robust subset of SNPs.

##### Stable Set Construction

Finally, the whole procedure (i.e. SNP ranking and subset selection) is repeated multiple times to obtain stable set of features. For an illustrative example, let’s assume we run the entire procedure *p* times. Consequently, we get *p* rankings of SNPs and *p* subsets of robust SNPs (through ranking and subset selection procedure). Finally, we extract common SNPs among the *p* subsets. Suppose, *t*_*i*_ represent a robust subset of SNPs from *i*^*th*^ repetition where 1 ≤ *i* ≤ *p*. Then the stable subset of SNPs will be *T* = *t*_1_ ∩ *t*_2_ ∩ *t*_3_ ∩ … ∩ *t*_*p*_.

##### Cross Validation

Cross validation splits data into a training, or discovery, set *D* used to determine a set of SNPs *P*_*D*_ that passes a threshold predicting a phenotype, and a test set *V* independent of the training set, used to test, or validate, the predictive power of the test *P*_*V*_ on the training set’s best variants. This is used in the “Feature Set Selection” and “Stable Set Construction” stages of our nested training-test set configuration. Therefore, a major component of power analysis of these methods revolve around evaluating factors impacting cross validation.

The discovery set parameters are ℙ (*P*_*D*_|*Ā*) = *α*_*D*_ and ℙ_D_(*P*_*D*_|*A*) = 1 − *β*_*D*_, and the validation set parameters are ℙ (*P*_*V*_ |*Ā*) = *α*_*V*_ and ℙ_𝕍_(*P* |*A*) = 1 − *β*_*V*_. ℙ (*A*) = *f* in both groups. *P*_*D*_ and *P*_*V*_ are assumed to be independent. That is ℙ (*P*_*V*_ |*P*_*D*_) = ℙ (*P*_*V*_), etc, and ℙ (*P*_*V*_ ∩ *A*|*P*_*D*_ ∩ *A*) = ℙ (*P*_*V*_ | ∩ *A*), Following arguments above,

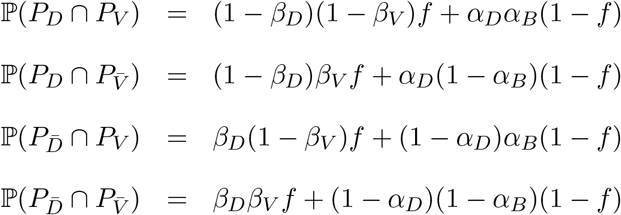

These relations satisfy 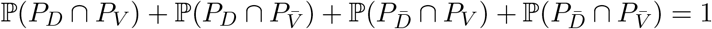.

We apply the above to estimate numbers of SNPs expected to be shared between training and test sets in some of the steps of our ML algorithm.

An odds ratio may be defined

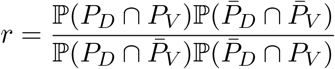

If there were no biologically active SNPs, then *f* = 0, and *r* = 1. So this can test whether the number of SNPs in both test and training sets is more than expected by chance.

## Model selection

We use two different ML algorithms for the two steps outlined in the RubricOE pipeline (Figure 2), IFS and SSC. We use Ridge Regression for ranking the features in IFS and LSVM to score the Youden curves for optimization.

One standard option for selecting the initial feature ranking has been to use regression coefficients. In our approach, one method for ranking in the ‘IFS stage is ridge regression ranking by magnitude of the coefficients. Another alternative was to consider the variation in the phenotype covered by the coefficient.

In standard regression, given cov(*y, y*^*T*^) = Σ, a *χ*^2^ distributed statistic measuring goodness of fit of a model *y* = *Xb* is

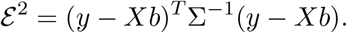

This may be rewritten

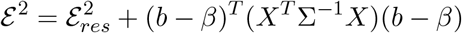

Where

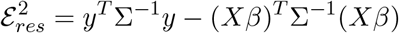

with

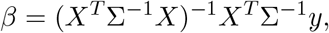

and

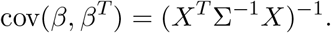

Given this, it is notable that the residual may be written

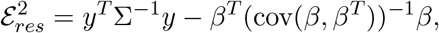

noting that *X* and *b* were augmented by a feature identical to unity in *X*, and equal to an offset in *b*. This gives a measure of variability to the variabiity in the mean for *y*. Therefore, the multivariate equivalent of sums of squares of the z-scores, including effects of correlations, represents the contribution of the predicted values to the residual. We identify the unexplained variability to be 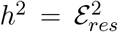, with the proportion explained by individual features 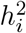 for *β*_*i*_ as 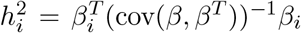 for each component by itself. We note that *y*^*T*^ Σ^*−*1^*y* is not centered. The contribution from offsets to the variability are included in the offset variable included in the augmented *β*’s offset estimation. Centering *y* shifts the offset, but does not change the coefficients. The residual error 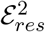 represents the proportion of phenotype not predicted by the regression model, including genetics and adjustment.

Feature selection regressions tend to be underdetermined, so a regularization that preserves the form of 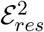, and that provides error bars and covariances for the coefficients is desirable. This argues for an *L*2 regularization. In this case, it is assumed that the coefficients *b* in the regression are themselves distributed, and have terms with variance *C*^*−*1^ and mean 0. Then for cov(*y, y*^*T*^) = Σ, and cov(*b, b*^*T*^) = *C*^*−*1^*I* for a number *C*, a *χ*^2^ distributed statistic measuring goodness of fit of a model *y* = *Xb* is

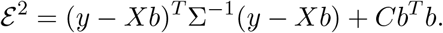

This may be rewritten

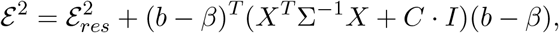

where

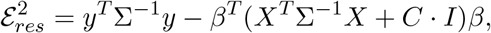

with

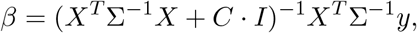

and

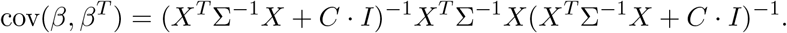

In the above, the

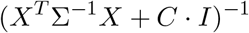

term induced by regularization tempers the response of *β* to variations in *y*. But the contribution to 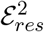 that reflects the cov(*β, β*^*T*^) contribution is the (*X*^*T*^ Σ^*−*1^*X*), while the *C* · *I* does not reflect predictive information. Expanding, this becomes

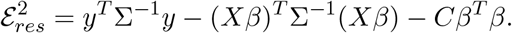

Given this, note that

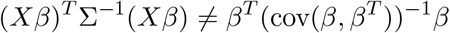

So the amount of variation due to the y variation accounted for in the residual is not equivalent to the z-score squared of the coefficients in the way that it is in non-regularized linear regression. Note that the part of the contribution to 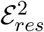 that responds to *y* is (*Xβ*)^*T*^ Σ^*−*1^(*Xβ*). However, while the direct relationship between variability is lost in ridge regression, the expression capturing the dependency on *y* variation should be applied for ranking, using the 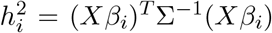 as in standard regression described above, but with the ridge-regression estimated coefficients. Note also that *Xβ* is still the estimator for *y*, so (*Xβ*)^*T*^ Σ^*−*1^(*Xβ*) is still the component that describes contributions to the proportion of variability in *y*^*T*^ Σ^*−*1^*y*. We apply ridge regression to feature selection as described above, computing the regression in the singular-value decomposition (SVD) basis. The value of the regularization parameter *C* is chosen to be the median singular value obtained from the SVD.

### Characterizing Stable Features

Logistic regression is evaluated for the stable set applied to the training sets and to the final test set to understand how cross-validated selection of final features relates to their estimated uncertainties, and to understand more clearly the power analysis situation in this problem.

### Code Availability

RubricOE executable is available at https://github.com/ComputationalGenomics/RubricOE.

## Data Availability

All simulated data was used in this manuscript.

## Data Availability

All data were simulated using the simulator described in https://www.nature.com/articles/ng.3244

## Acknowledgements

This work was conducted by SS as part of his Postdoctoral Research at IBM T.J Watson Research Center.

## Appendix

### Simulated Data

**Figure 3:**
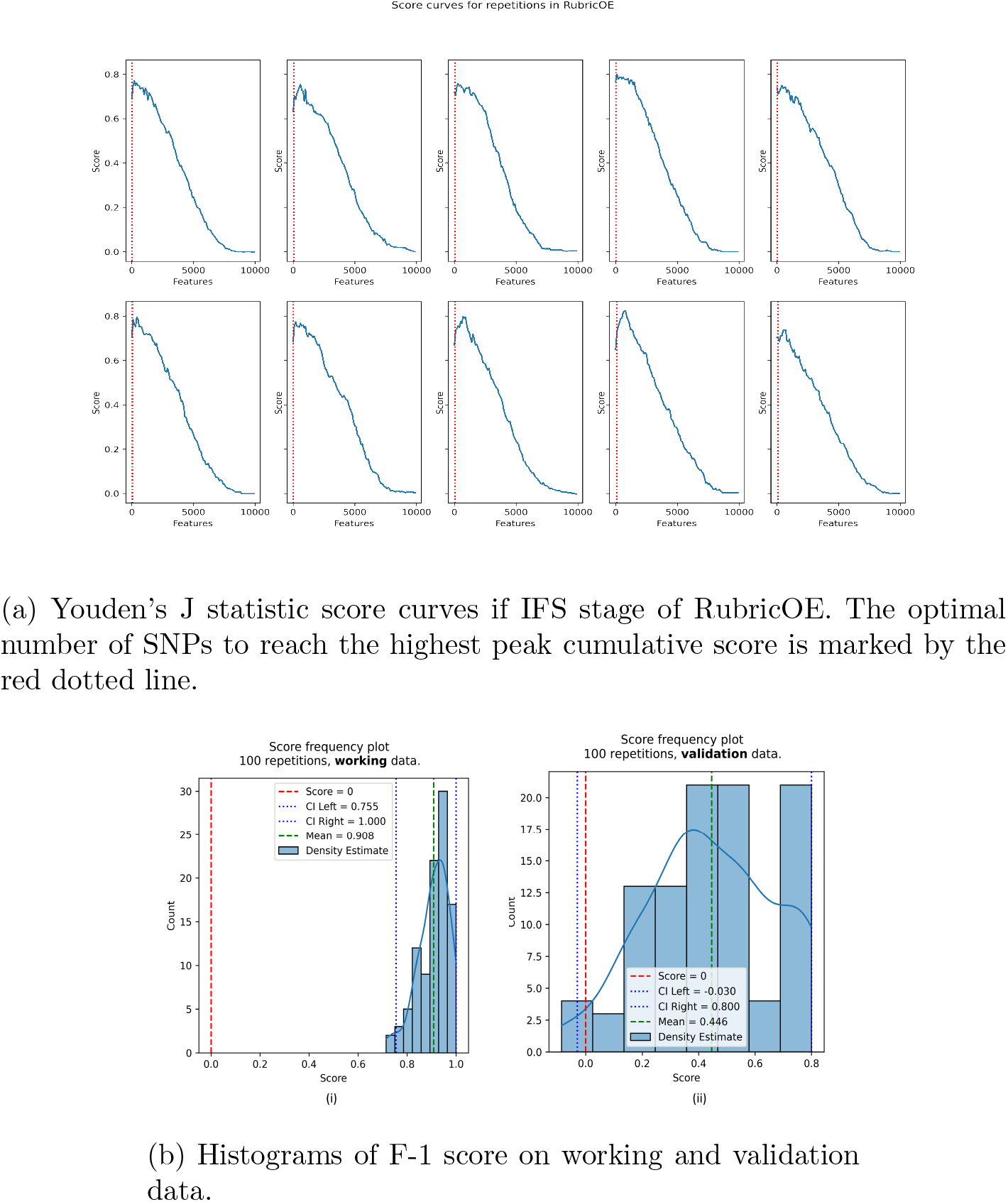
Performance of RubricOE on PSD scenario with 70% genetic effect.

**Figure 4:**
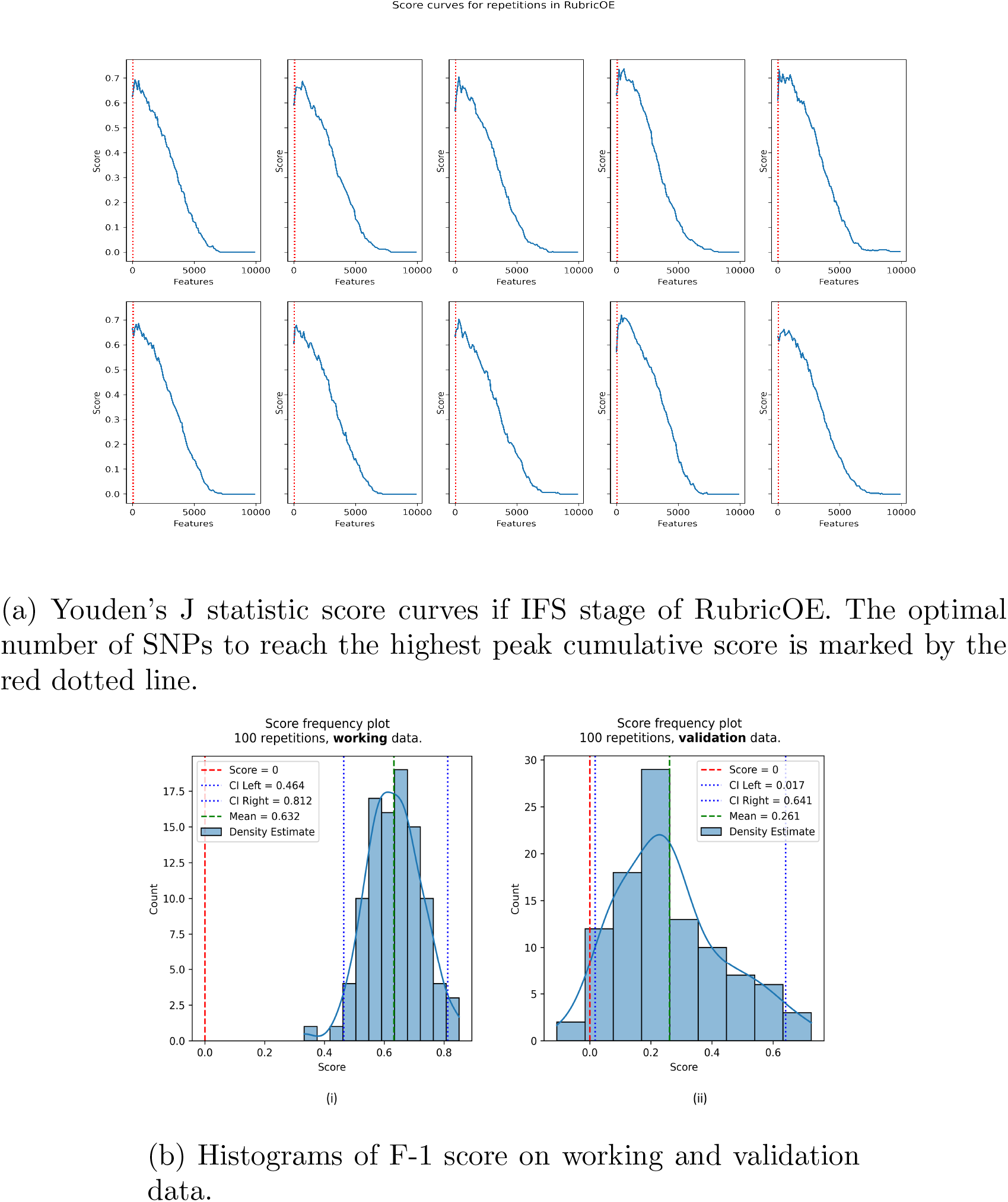
Performance of RubricOE on TGP scenario with 70% genetic effect.

**Figure 5:**
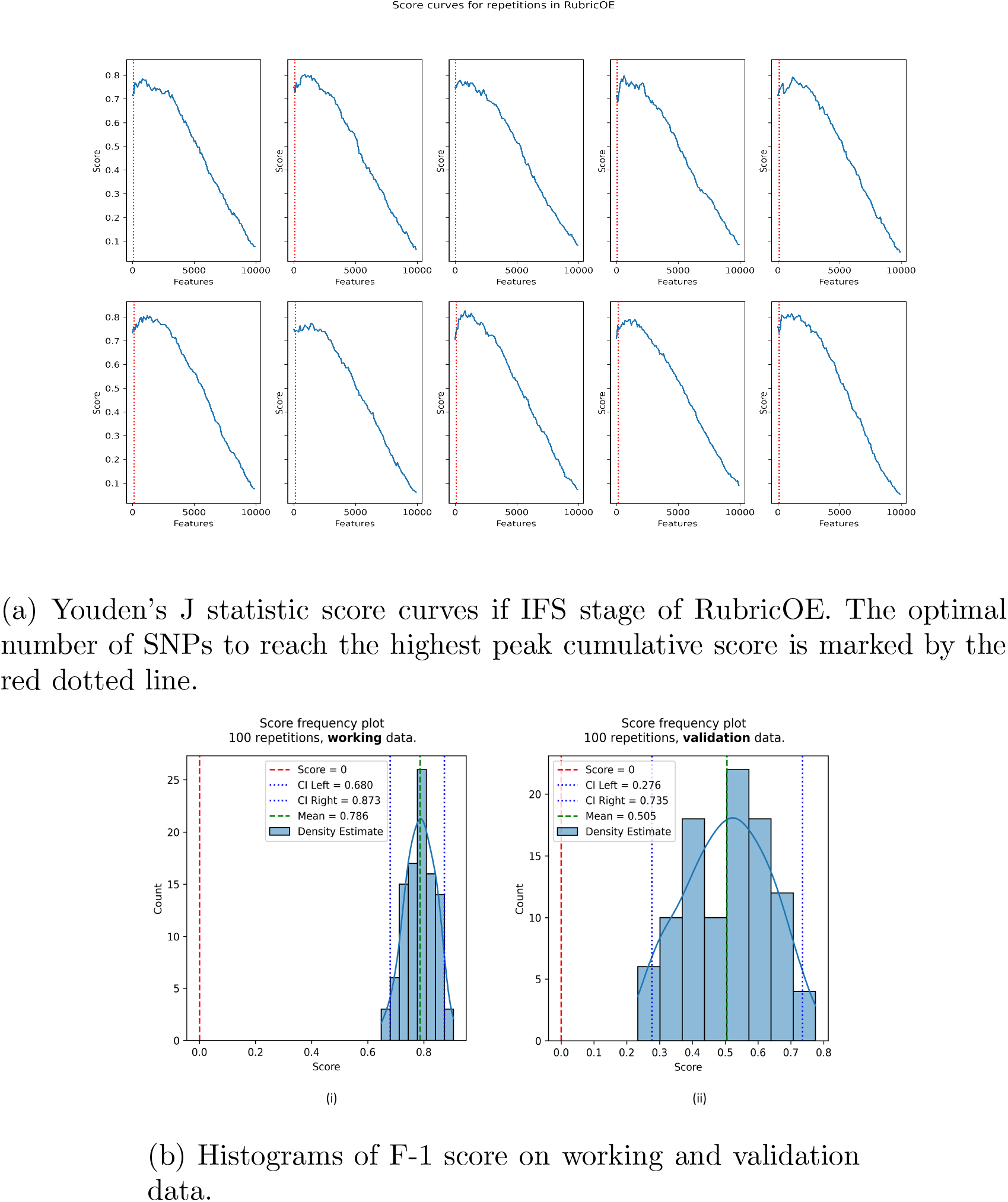
Performance of RubricOE on BN scenario with 70% genetic effect.

